# Contraceptive Adoption and Continuation among Postpartum Women in India’s urban slums

**DOI:** 10.1101/2025.08.01.25332810

**Authors:** Emily Das, Mukesh Kumar Sharma, AJ Francis Zavier, Hitesh Sahni, Ian Salas, Kojo Lokko, Kate Graham, Samarendra Behera, Neelanjana Pandey, Jessica Mirano

## Abstract

The unmet need for postpartum contraception provides a critical opportunity to introduce modern contraceptives due to the increased use of maternal care health services. However, limited knowledge exists regarding program reach in urban slum settlements and the impact of interventions on improving the adoption and continuation of modern contraceptives. The study aimed to assess the influence of health worker exposure and service delivery point visitation on contraceptive use within the postpartum period.

This study utilized data from a subset of an output tracking survey conducted in 2019, focusing on 1,503 women aged 15-34 years who had given birth within the last three years. Multinomial logistic regression and proportional Hazard models were employed to assess the program’s effect on contraceptive adoption and continuation. Among urban slum residents, 38% to 46% initiated the use of modern reversible contraceptives within six and 12 months after delivery. Interpersonal communication with ASHA/FPA (Accredited Social Health Activist /Field Program Associates) inter-spousal communication, and self-efficacy were significant predictors of contraceptive adoption, with the latter two variables remaining significant at 12 months. Women with higher parity, education, and self-efficacy were less likely to discontinue the adopted method.

While program exposure influenced contraceptive adoption, the analysis did not find an impact of health worker interaction or service delivery point visitation on the continuation of postpartum contraception. This study emphasizes the need for targeted interventions and continuous support to improve adoption and continuation rates. Barriers such as inter-spousal communication and self-efficacy should be addressed to enhance the effectiveness of postpartum family planning programs. Further research is needed to develop comprehensive strategies that address these challenges and improve postpartum contraceptive outcomes.

## Introduction

Promoting safe sexual and reproductive choices and voluntary family planning is a global commitment aimed at achieving sustainable development goals, to enhance maternal health, and reduce the unmet need for contraception by 2030 (Abraha et al., 2017; Ndugwa et al., 2011; Singh et al., 2014; Rana & Goli, 2021). Family planning plays a pivotal role in enabling individuals and couples to attain their desired number of children and appropriately space their births, thereby leading to reduced rates of maternal and infant mortality (Vernon, 2009; World Health Organization & Reproductive Health, 2007; UNFPA, 2020). Timely and effective access to postpartum contraception is crucial for improving women’s well-being during the postpartum period (Tazneen et al., 2017) and reducing adverse health outcomes such as maternal anemia, low birth weight, and high neonatal/infant mortality (Rutaremwa et al., 2015; Dixit et al., 2017).

However, the utilization of postpartum contraception remains low, particularly in low- and middle-income countries like India (Moore et al., 2015; Shah et al., 2015; Singh et al., 2014; Achyut, 2016). Barriers such as limited awareness, cultural attitudes, inadequate access to facilities, and insufficient counseling contribute to the unmet need for contraception during the postpartum period (Singh et al., 2014; Thapa et al., 1992; Achyut, 2016). A systematic review conducted by Cleland et al. (2015) on interventions to improve postpartum family planning suggests that counseling before discharge and integration of family planning into immunization and pediatric services could serve as opportunities to promote the use of postpartum family planning. Consequently, there is an urgent need to implement targeted programs that address these barriers and enhance the uptake of postpartum contraception among women in India.

More than 257 million women in low- and middle-income countries who desire to delay childbearing are not utilizing any form of modern contraception (Tazneen et al., 2017; UNFPA, 2020). Numerous studies have demonstrated that women often exhibit inconsistent use of postpartum family planning methods, resulting in unintended pregnancies, and high rates of contraceptive discontinuation during the postpartum period (Ali, Cleland, & Shah, 2012; Warren, Abuya, & Askew, 2013; Kopp et al., 2017; Mumah et al., 2015). Furthermore, the lack of contraceptive use and unmet need is particularly prevalent among specific population clusters, including urban slum residents in low- and middle-income countries (Mody et al., 2014; World Health Organization, 2018; Coomson & Manu, 2019; Roy et al., 2021). However, there has been limited research on contraceptive continuation and switching during the postpartum period in low-resource settings. Studies suggest that several barriers hinder women from accessing postpartum contraception, such as lack of awareness, inadequate inter-spousal communication, cultural attitudes, limited agency among women, inadequate facility access, and insufficient counseling skills among healthcare providers (Salway & Nurani, 1998; Barman, 2013). However, studies have also indicated that women in the postpartum period prefer longer birth intervals and would be more inclined to use contraception earlier if they had access to better information regarding their risk of pregnancy after childbirth or if they were provided with a wider range of options to achieve their desired birth spacing (Pasha, 2015). Moreover, research has demonstrated that women with higher agency are more likely to utilize postpartum contraception (Keerara et al., 2017; Smith et al., 2017; Willcox et al. 2021). A recent systematic review conducted by Robinet et al. (2023) underscored the significance of women’s decision-making in the context of postpartum contraceptive use. Additionally, the lack of spousal communication or support has been identified as a barrier for women in adopting postpartum contraception (Jennifer, 2021). In lower and middle-income countries, spousal communication regarding reproductive health is often insufficient (Santhya and Jejeebhoy, 2015). Notably, a community-based intervention study conducted in India revealed that a 43 percent increase in spousal communication resulted in a 27 percent increase in the utilization of contraception among young married women (Behera et al., 2016).

Counseling for postpartum contraception has been found to increase its adoption (Puri et al., 2021b; Puri et al., 2023; Asah-Opoku, 2023; Srivastava et al., 2022; Zavier and Santhya, 2013), and providing family planning messages to pregnant women during their health facility visits increases the likelihood of adopting a modern method after childbirth (Veron, R., 2009; Speizer IS., 2013). Community-based programs have also proven effective in increasing the use of contraceptive methods during the postpartum period (Ahmed et al., 2013). A systematic review of postpartum contraceptive use suggested that six interventions showed some association with the acceptance of contraception during the postpartum period (Lopez et al., 2014). Recent evidence suggested that women value contraceptive counseling both during the antenatal and postnatal periods, highlighting the importance of providing comprehensive support (Freeman-Spratt et al., 2023).

India continues to face significant challenges in family planning, despite implementing a longstanding national program. The country grapples with a high unmet need for contraception and elevated rates of maternal morbidity and mortality. Although there has been improvement in the contraceptive prevalence rate among married women aged 15-49, disparities persist across states, rural-urban populations, and socioeconomic status (NFHS, 2015-16; 2019-21; Srivastava et al., 2022). Unintended births, miscarriages, and induced abortions remain prevalent, underscoring the necessity for effective family planning efforts (Singh et al., 2018). Furthermore, during the postpartum period, women in developing countries, including India, often engage in sexual relationships without contraception, leading to an increased risk of unintended pregnancies (Rajan et al., 2016). The launch of the Janani Suraksha Yojana, a conditional cash transfer program promoting institutional deliveries, provides an opportunity to promote postpartum contraception (Zavier and Santhya, 2013).

Urbanization is rapidly increasing in India, with approximately 34% of the population residing in urban areas, including a significant number of impoverished urban residents (Sethi, 2020; Hazarika, 2010; Achyut et al., 2016). Urban slums are characterized by various challenges, such as high population densities, poverty, inadequate housing, poor hygiene, and limited access to essential services, including sexual and reproductive health rights (Ndugwa et al., 2011; Rutaremwa et al., 2015; Rajan et al., 2016; Achyut et al., 2016; Khan & Kotecha, 2019). This study aims to provide evidence on contraceptive use among postpartum women in urban slums of India and explore how interventions and programs can improve contraceptive adoption. The study examines the levels, adoption rates, and continuation rates of postpartum contraception among women in urban slums, as well as the potential influence of health worker outreach and program interventions.

### The Program Intervention

Population Services International (PSI), in partnership with the National Health Mission (NHM) and with support from the Bill & Melinda Gates Institute for Population and Reproductive Health, and USAID has implemented a project entitled “The Challenge Initiative for Healthy Cities (TCIHC)” to strengthen existing service delivery platforms at the city-level health systems to improve demand for and access to family planning services for urban poor in India. The TCIHC program was implemented between 2016 and 2020 in 31 cities of India across three states, namely – Uttar Pradesh, Madhya Pradesh, and Odisha.

Under the program, the PSI helped the NHM to create coaching platforms across all levels of health administration – state, division, district, block, and community levels. The coaching focused on improving the ability of healthcare providers, primarily Accredited Social Health Activists (ASHA) to address the issues/challenges faced by the family planning programs in the respective cities. As part of the TCIHC coaching intervention, the ASHAs were provided with printed handouts of training material along with other job aids to help them improve their ability to counsel eligible couples. ASHAs were also coached on various key issues such as maintaining the urban health index register (UHIR), the basket of choices, and the male engagement strategy. The ASHAs were also trained to have more confidence while counseling couples to adopt suitable contraceptive methods, especially among first-time parents.

The facility-based TCIHC program focused on ensuring the optimal family planning service provision at health facilities such as urban primary health centers (UPHC), outreach camps (ORC), and urban health and nutrition days (UHND). Under the program site orientations were conducted at these facilities/health service delivery points to ensure that these sites can effectively deliver activities aimed at improving knowledge and information about family planning, widespread availability and access to family planning services, and uptake in contraceptive use. For example, the provision of Adolescent and Youth Sexual and Reproductive Health (AYSRH) services at the UPHC level to ensure that all staff is trained to facilitate reproductive health counseling for adolescents on 2-3 select days a week. Additionally, the TCIHC program also helped coach UPHC staff on new contraceptive options introduced in the Indian family planning program, namely Antara (injectables) and Chayya (pills). This included training on these newer contraceptives and their potential side effects, along with effective management of supply and other logistics. Hence, the entire coaching intervention of the TCIHC is broadly categorized as follows – (i) improving the capacities of Accredited Social Health Activists (ASHAs) and other frontline health workers, (ii) ensuring optimal functioning of UPHC, UHND, and ORC which are, usually the first point of contact for a community member within the urban public health system, and (iii) enabling data-based planning of activities.

### Data

As part of a program intervention, two rounds of surveys were conducted in 14 cities across three states in India. The surveys, known as Output Track Surveys (OTSs), were conducted among currently married women aged 15-49 years in both slum and non-slum areas. Individual data was collected from a representative sample of 8,319 women during the second round of OTS in September 2019, with 4,194 women surveyed in slum areas and 4,125 in non-slum areas. The focus of analysis for this paper was the data collected from slum areas, which were identified in each city through a mapping and listing process with local officials. The analysis specifically looked at data from 1,503 women aged 15-34 years who had given birth within the last three years before the survey.

### Ethical considerations

Before the survey was conducted, approvals were granted by the Institutional Review Board at Johns Hopkins Bloomberg School of Public Health (IRB Number-8412), Population Services International, and locally by SIGMA-IRB (CIN No: U74140DL2008PTC182567) and are consistent with international standards for the ethical conduct of research. Participation was voluntary and confidential for all respondents. Informed consent was obtained from all respondents before the interview, either in writing or orally.

## Variables

### Dependent variables

1) Use of contraception during the postpartum period, that is, within three months, six months, and 12 months, separately. For measuring the adoption of postpartum contraception, women who had a delivery at least three months ago were included in the analysis. Similarly, contraceptive use within 12 months of the postpartum period includes those women who had delivered a child for more than 12 months at the time of the survey. These two variables were coded as 1) did not adopt any method, 2) adopted permanent methods, 3) adopted modern reversible methods, and 4) adopted traditional methods.

2) Number of months of use of contraception after birth for the analysis of postpartum contraceptive continuation rate. The contraceptive use status variable is defined as one if women discontinued the adopted method and zero if they continued at the time of the survey.

### Explanatory variables

The main explanatory variables used were program exposure variables 1) exposure to family planning information through health workers - if the woman met ASHA (Accredited Social Health Activist) or with FPA (Field Program Associates) for family planning information and counseling in the last six months before the survey, and 2) exposure to family planning information at service delivery points - if a woman visited a service delivery point such as UPHC (Urban Primary Health Centre), ORC (Outreach Camp) or UHND (Urban Health and Nutrition Day) to seek family planning services in six months before the survey.

Other independent variables included in the analysis encompassed age, education, number of previous children, religion, caste, employment status, wealth index, spousal communication on family planning, exposure to family planning messages via television, the self-efficacy index, and the index of decision-making about the use of family planning. A qualitative review informed the selection of these background variables of 34 studies conducted in India and abroad on the use of contraception during the postpartum period (Robinet et al., 2023). The inter-spousal communication refers to spousal communication regarding family planning in the last 3 months. Most of the other variables are self-explanatory, except for the wealth index, self-efficacy index, and index of decision-making about the use of family planning. Detailed information regarding the calculation of the wealth index can be found in Appendix Table 1.

**Table 1:**
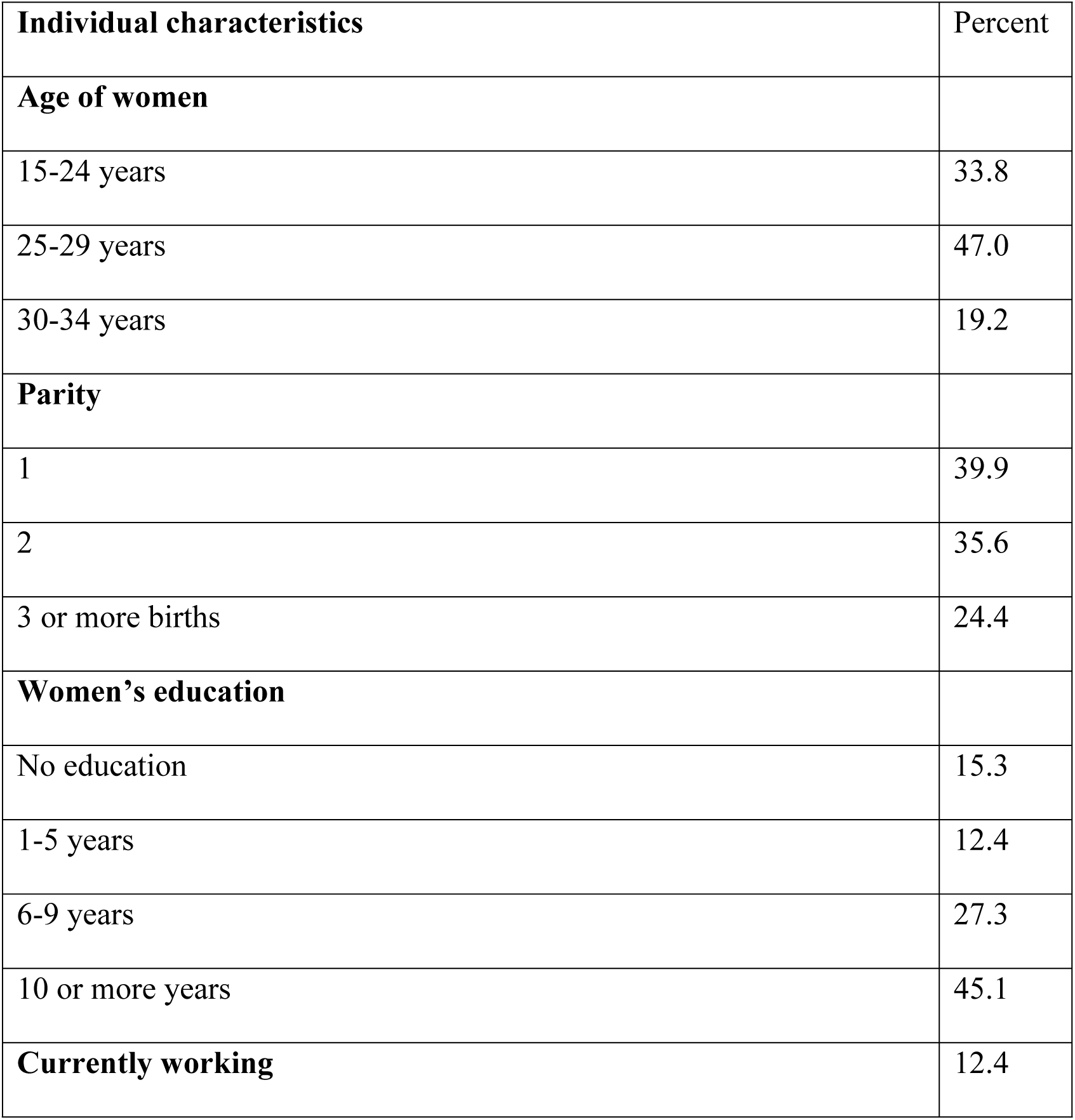
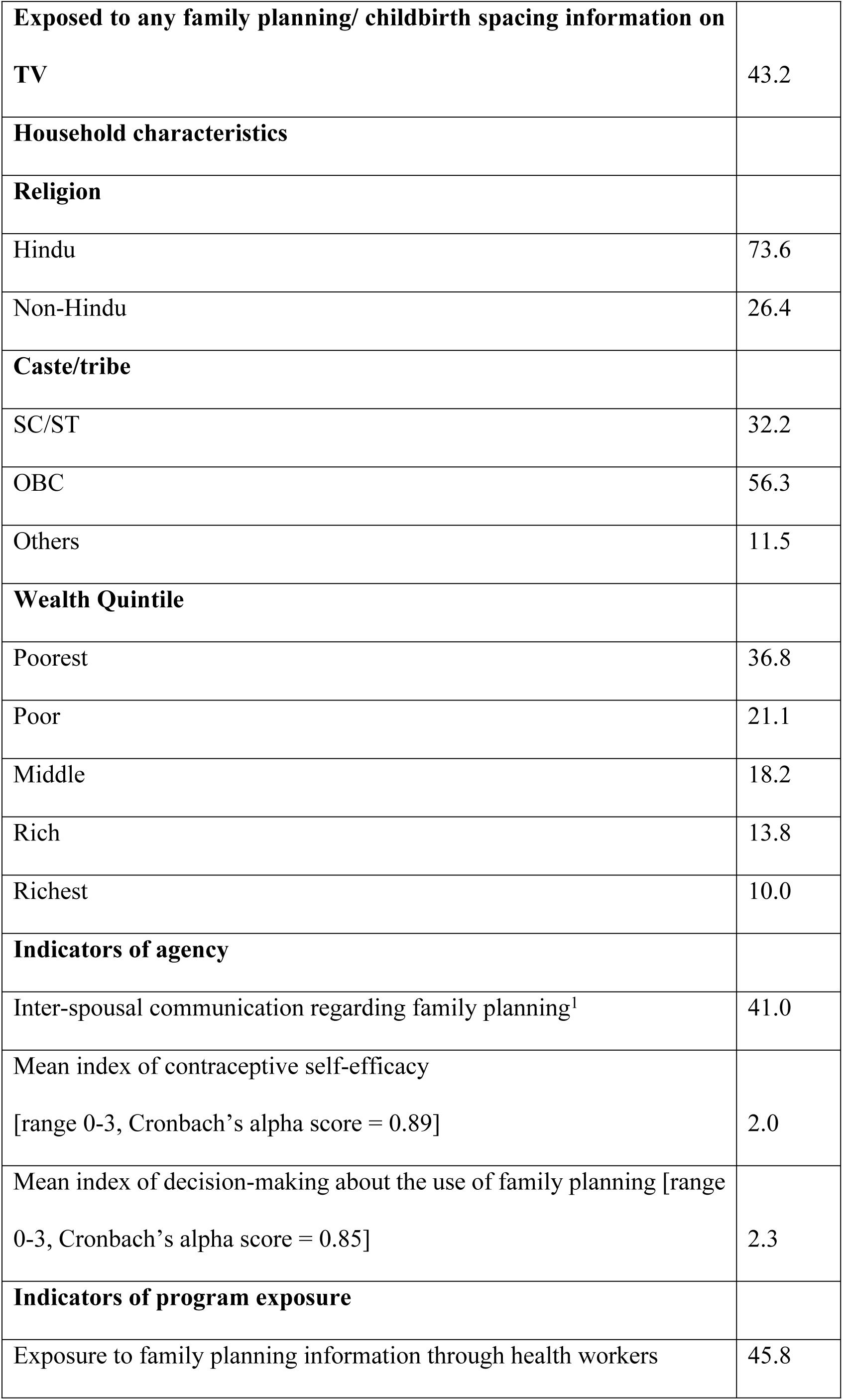
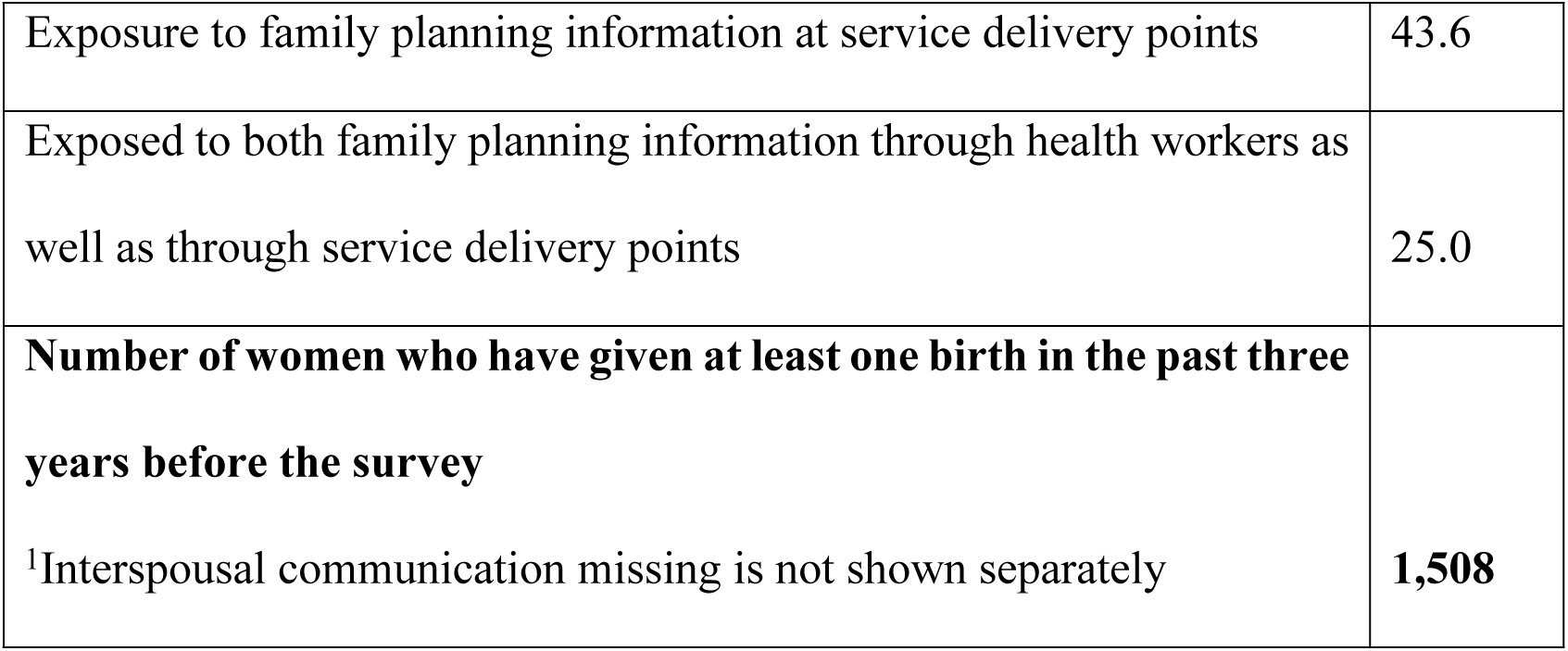
Profile of the women aged 15-34 years living in slums of TCIHC Cities, 2019

The index of contraceptive self-efficacy was derived from three statements, with responses ranging from one to five, indicating the degree of agreement or disagreement, varying from strongly disagree to strongly agree. The three statements employed were as follows: 1) I am capable of correctly utilizing the family planning method of my choice, 2) I am able to consistently employ the family planning methods of my choice, and 3) I feel confident in my ability to obtain a family planning method. The responses of “somewhat agree” and “strongly agree” for all three statements were assigned a code of 1, while all other responses were coded as 0. The Cronbach’s alpha coefficient for these three variables was determined to be 0.89. The index of contraceptive self-efficacy was calculated by summing the values of all three variables, resulting in an index ranging from zero, indicating low contraceptive self-efficacy, to three, indicating high contraceptive self-efficacy.

Similarly, the measurement of the index of decision-making about the use of family planning was conducted using three variables: 1) determining who possesses the predominant influence over the usage of a contraceptive method, 2) ascertaining who possesses the primary authority over the choice of contraceptive method, and 3) identifying who holds the primary decision-making power regarding the timing of having a child. The responses to each of these statements were categorized into five options: respondent alone, the husband alone, respondent and husband equally, respondent and someone else, and someone else. The responses of “respondent alone” and “respondent and husband equally” for all three statements were assigned a code of one, while all other responses received a code of zero. The index of decision-making about the use of family planning was computed by summing these three variables. The index measures women’s say in the use of contraception and having a child. The Cronbach’s alpha coefficient was determined to be 0.85, and the resulting index ranged from zero indicating a low level of decision-making ability, to three, representing a high level of decision-making ability.

## Statistical Analysis

Descriptive analyses were undertaken to examine the attributes of women and the extent of contraceptive usage. Bivariate analyses were conducted to evaluate the associations between the variables. Additionally, multinomial logistic regression analysis was employed to demonstrate the impact of the program on the adoption of contraception during the postpartum period. The adoption of postpartum contraception is categorized into four groups: those who did not adopt a method, those who adopted permanent methods, those who adopted modern reversible methods, and those who adopted traditional methods. To ascertain the continuation of postpartum contraception, Kaplan-Meier survival curves and a proportional hazard model were utilized. The Kaplan-Meier survival curves display the unadjusted continuation rate of contraception. The Proportional Hazard Model is a statistical model employed in survival analysis to investigate the relationship between independent variables and the hazard rate, which represents the risk of an event occurring over time. This model offers valuable insights into the factors influencing survival outcomes or the time it takes for an event to occur.

## Results

### Profile of sample women

Young women between the ages of 15 and 24 constituted approximately one-third (34%) of the sample. Among the sample women, almost 40% were experiencing their first pregnancy, while 36% were in their second pregnancy, and approximately a quarter of the women had higher parity levels (as shown in Table 1). Around 45% of the women had received an education for 10 or more years, while 15% had not attended school. At the time of the survey, only a small proportion of women (12%) were engaged in employment. Roughly 43% of the women had been exposed to family planning or childbirth spacing information through television. The majority of respondents (74%) identified as Hindus, and about one-third (32%) belonged to scheduled castes and tribes, which are considered socially disadvantaged groups. More than half (58%) of the women hailed from households categorized as poor or falling under the poorest bracket in terms of household wealth.

In terms of interspousal communication, approximately 41% of women reported engaging in such discussions. The average score for the family planning self-efficacy index was two out of three items, and the mean score for the decision-making index regarding the use of family planning was 2.3 out of three.

Around 46% of women had received family planning information from healthcare workers, and 44% had encountered such information at service delivery points.

### Topics discussed during health workers’ visits

Women who had interacted with health workers were further asked about the topics discussed during the visits. Table 2 details the topics covered during these interactions. More than two-fifths of the women reported that health workers explained the need for using family planning methods (46%) and informed them about the various family planning options available (40%). However, only 17% of the women were informed about the possible side effects of each method. Additionally, 11% of the women mentioned that health workers supported them in making family planning decisions and choosing a method. A few women reported being informed about visiting a UHND (12%) or a UPHC (7%) to obtain family planning services

**Table 2.**
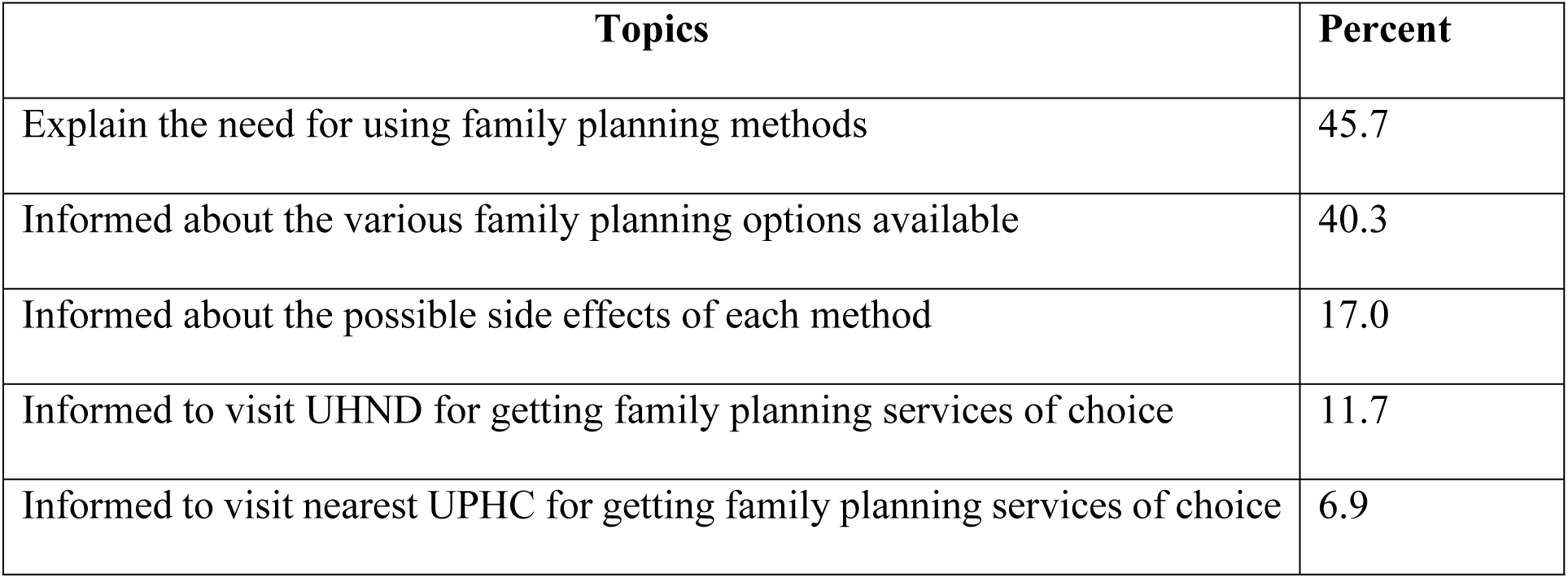
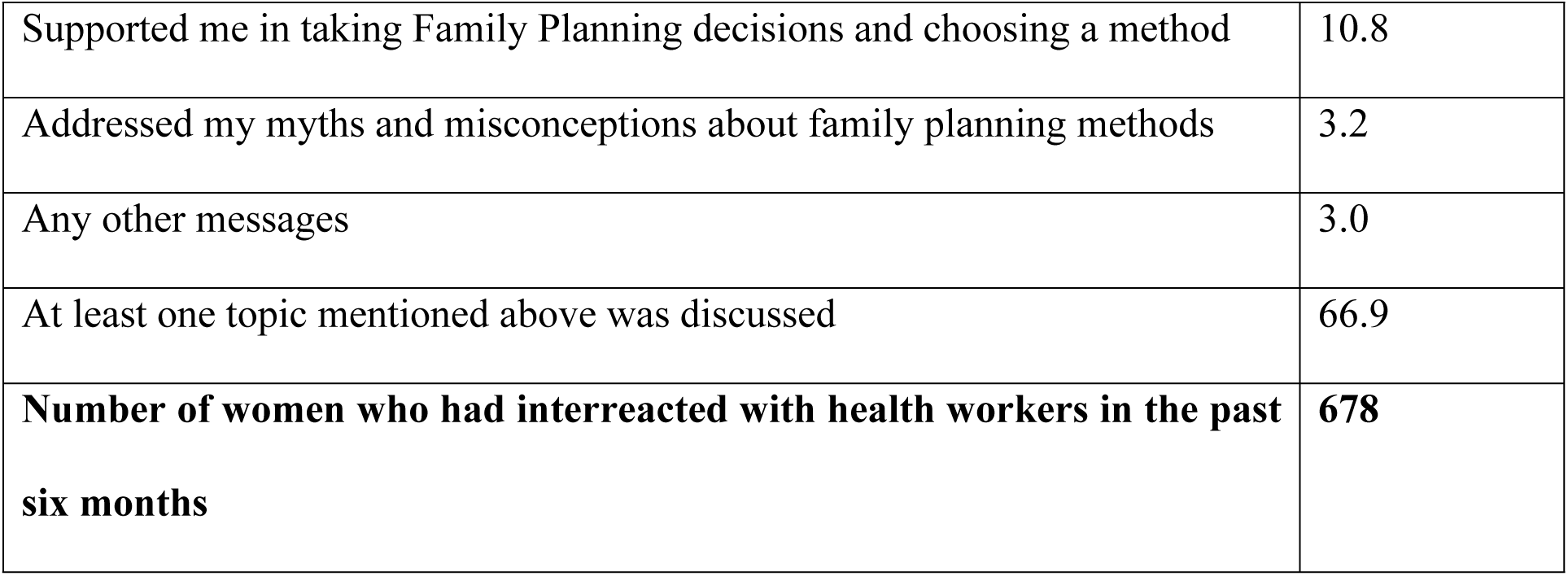
Percentage of women who met with health workers and topics discussed during visits

### Postpartum contraceptive use

Table 3 presents detailed information on postpartum contraceptive method adoption among the study population. Within the first six months following childbirth, approximately 56% of women who had given birth within the previous six to 36 months adopted some form of contraceptive method. Most of them opted for modern reversible methods (38%). Male condoms were the most adopted method (20%), followed by the intrauterine contraceptive device (IUCD) (10%). Traditional methods were reported by 12% of women. Additionally, within the first six months, eight percent of women chose a permanent contraceptive method. As the postpartum period extended to 12 months, an increased number of women adopted any form of contraception, reaching 70%. The use of condoms rose to 26%, remaining the most frequently used contraceptive method. Moreover, the utilization of traditional family planning methods increased to 16%.

**Table 3.** Percentage of women who adopted a family planning method within six and 12 months of the postpartum period

|  | **Within six months** | **Within 12 months** |
| --- | --- | --- |
| Not using any method | 42.5 | 29.9 |
| **Permanent methods** | **7.9** | **8.3** |
| Female sterilization | 7.4 | 7.8 |
| Male sterilization | 0.5 | 0.5 |
| **Modern reversible methods** | **37.5** | **45.5** |
| IUCD | 9.5 | 10.0 |
| Injectables | 1.8 | 2.4 |
| Daily pill | 1.3 | 2.1 |
| Weekly pill | 0.0 | 0.4 |
| Male condom | 19.6 | 25.5 |
| Spermicide/foam/jelly | 0.5 | 0.7 |
| Lactational Amenorrhea Method | 4.4 | 4.1 |
| Standard days method | 0.5 | 0.4 |
| **Traditional methods** | **12.0** | **16.2** |
| Rhythm method | 8.4 | 11.8 |
| Withdrawal | 3.6 | 4.4 |
| **Number of women who have given birth before six months / 12 months of the interview date in the last three years** | **1,097** | **820** |

### Exposure to the program and post-partum contraception

The data in Table 4 reveals that discussing family planning information with ASHA or FPA had a positive effect on post-partum contraception uptake. More women (61%) who talked to ASHA or FPA about family planning adopted a contraceptive method within six and 12 months compared to those who did not (53%). Similarly, a higher percentage (74%) of women who discussed family planning with ASHA or FPA adopted postpartum contraception within 12 months than those who did not (67%). There was no significant difference in the use of permanent and traditional methods between those who discussed family planning and those who did not. However, more women (43%) who discussed family planning with ASHA or FPA adopted modern reversible methods compared to those who did not (33%).

**Table 4.** Percentage of women who adopted a family planning method in the postpartum period by exposure to program

| **Adoption of postpartum contraception within 6 months of postpartum period** |  |  |  |  |
| --- | --- | --- | --- | --- |
|  | **Not using** | **Using permanent methods** | **Using modern reversible methods** | **Using traditional methods** |
| Exposure to family planning information through health workers [p=0.002] |  |  |  |  |
| No | 46.3 | 9.0 | 32.5 | 12.2 |
| Yes | 38.6 | 6.6 | 43.2 | 11.6 |
| Exposure to family planning information at service delivery points [p=0.214] |  |  |  |  |
| No | 42.2 | 7.4 | 37.9 | 12.5 |
| Yes | 43.3 | 8.4 | 37.0 | 11.4 |
| **Adoption of postpartum contraception within 12 months of the postpartum period** |  |  |  |  |
| Exposure to family planning information through health workers [p=0.025] |  |  |  |  |
| No | 33.0 | 8.7 | 40.9 | 17.3 |
| Yes | 26.5 | 7.8 | 51.0 | 14.8 |
| Exposure to family planning information at service delivery points [p=0.478] |  |  |  |  |
| No | 30.4 | 6.7 | 47.0 | 15.9 |
| Yes | 30.0 | 10.4 | 43.5 | 16.5 |

**Table 4.**
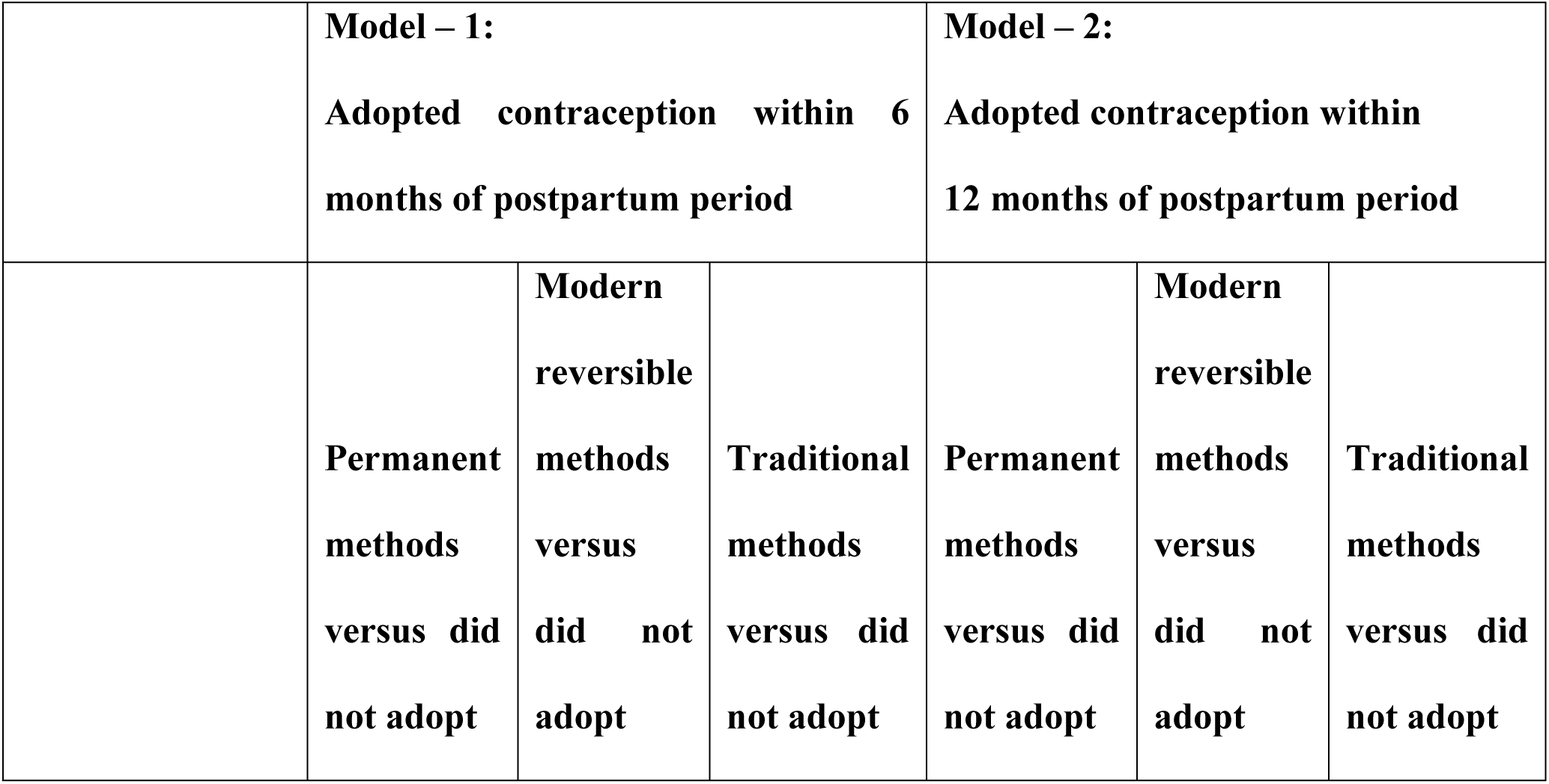
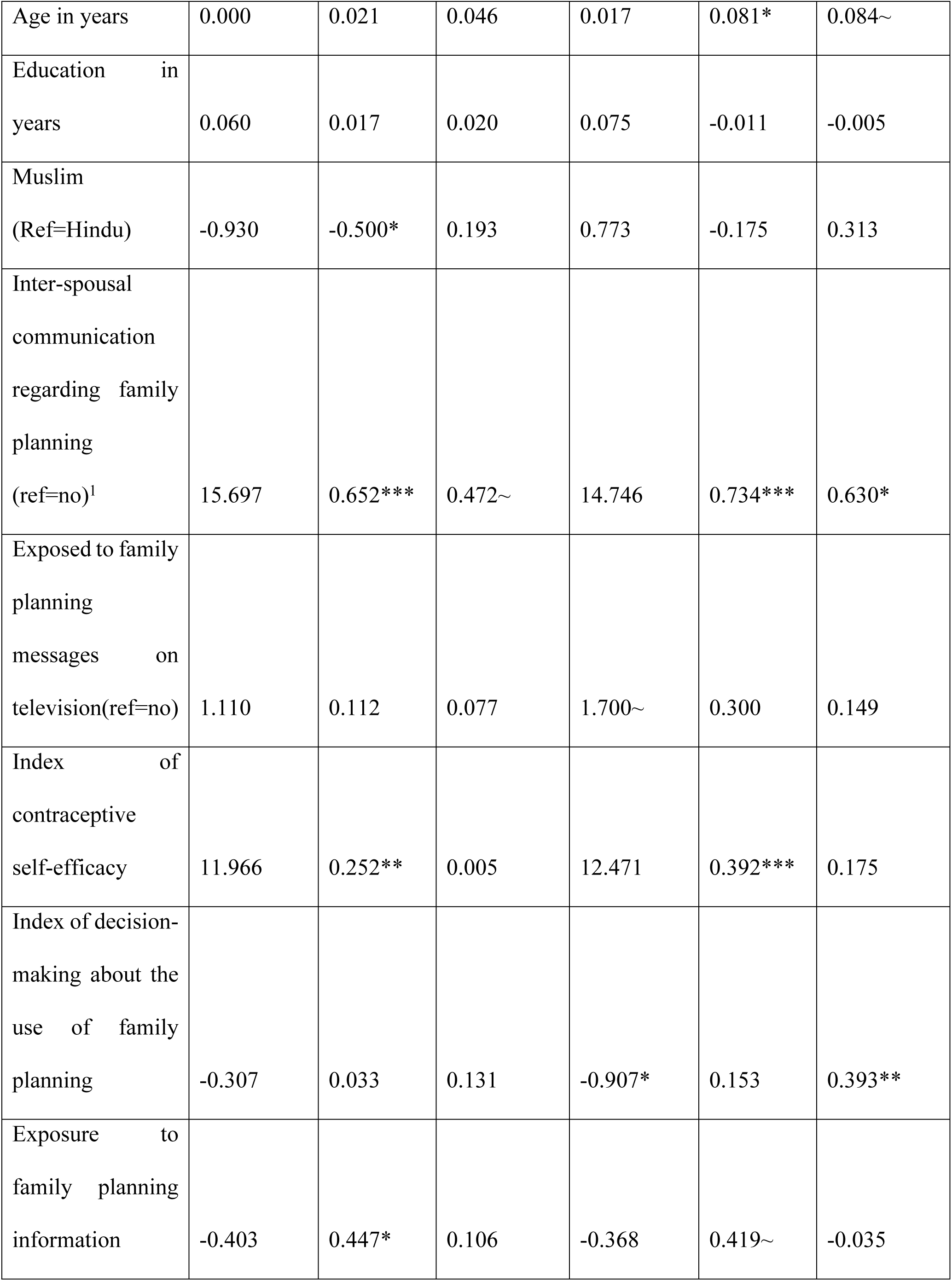
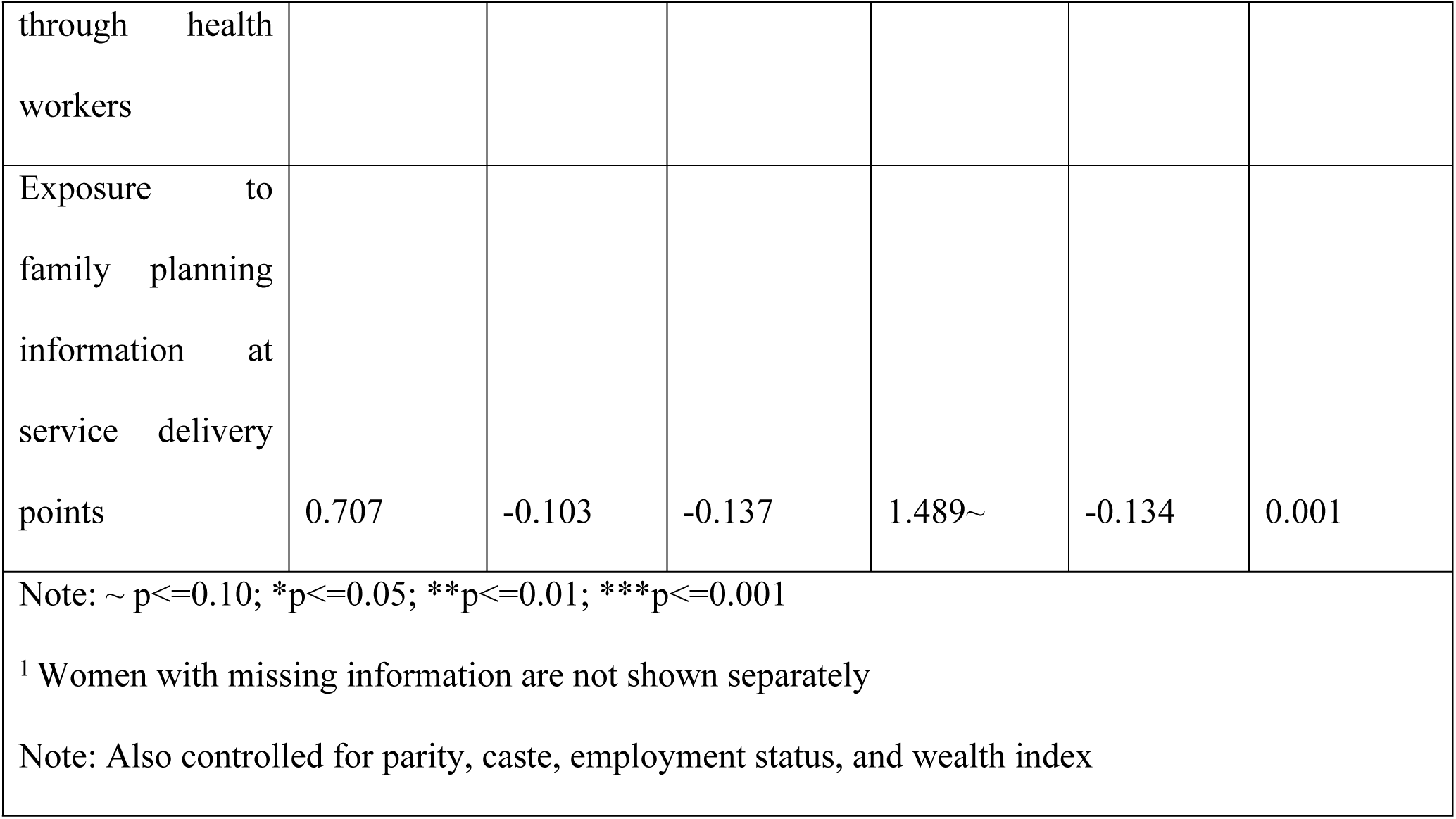
Coefficients of the multinomial logistic regression of postpartum contraception within six months and 12 months

Interestingly, exposure to family planning information at service delivery points did not have a noticeable impact on postpartum contraception uptake.

### Results from Multinomial regression analysis

Multinomial regression analysis was conducted to assess the impact of the program on postpartum contraceptive use within six and 12 months while controlling for relevant background and program exposure variables. The results can be found in Table 4.

The results indicate that the adoption of modern reversible methods is significantly associated with the program and other background variables when compared to women who did not adopt postpartum contraception. Among the two program variables, women who were exposed to family planning information through health workers (coefficient - 0.447, p<=0.050) were found to be statistically more likely to adopt modern reversible methods within six months of the postpartum period, in comparison to women without such exposure. Similarly, the association between exposure to family planning information through health workers and the adoption of contraception within 12 months of the postpartum period remained statistically significant, although the significance level was slightly weakened (Coefficient - 0.419, p<=0.100).

Furthermore, other variables that demonstrated significant associations with the adoption of modern reversible methods of contraception during the postpartum period included belonging to the Muslim religion, engaging in interspousal communication regarding family planning, and scoring higher on the self-efficacy index. Women who had discussions with their spouses regarding family planning were more likely to adopt reversible methods of postpartum contraception within both the six- and 12-month periods. Similarly, women with higher self-efficacy scores were also more likely to adopt reversible methods of postpartum contraception within six and 12 months after childbirth.

### Continuation of postpartum contraception

The continuation of postpartum contraception is explored using the KM survival curve. The continuation rate of reversible contraception at 12 months is more than 75% (Figure 1). Till 36 months, more than 50% of women continued the use of modern reversible methods. However, the KM survival curves estimated separately by program exposure did not differ statistically (Figures 2 and 3).

**Figure 1.**
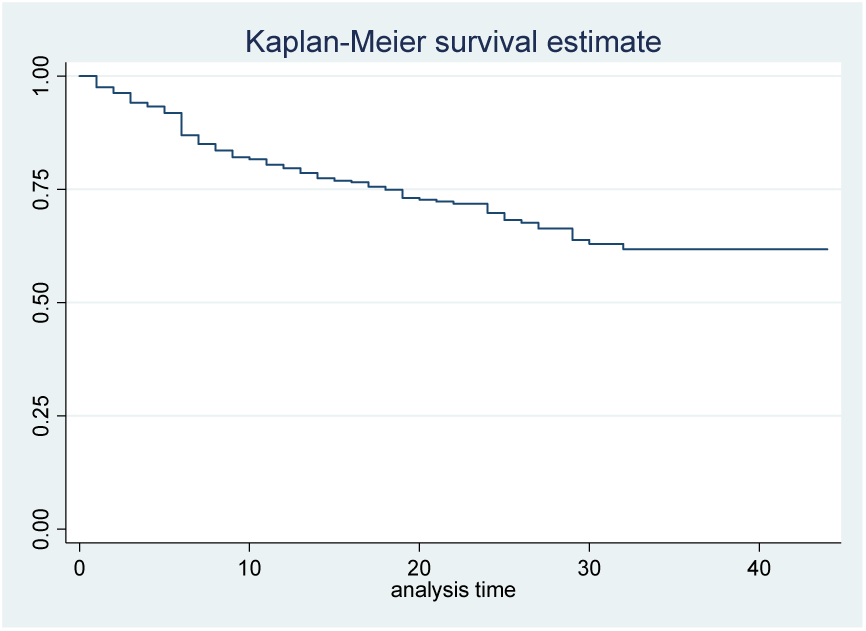
Continuation of postpartum modern reversible contraception

**Figure 2.**
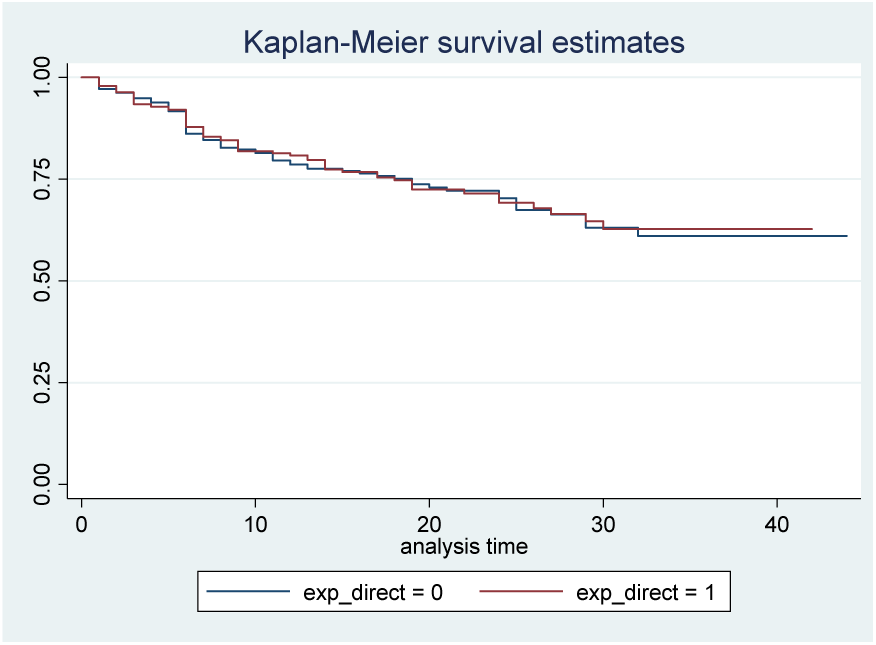
Continuation of postpartum modern reversible contraception by exposure to family planning information through health workers

**Figure 3.**
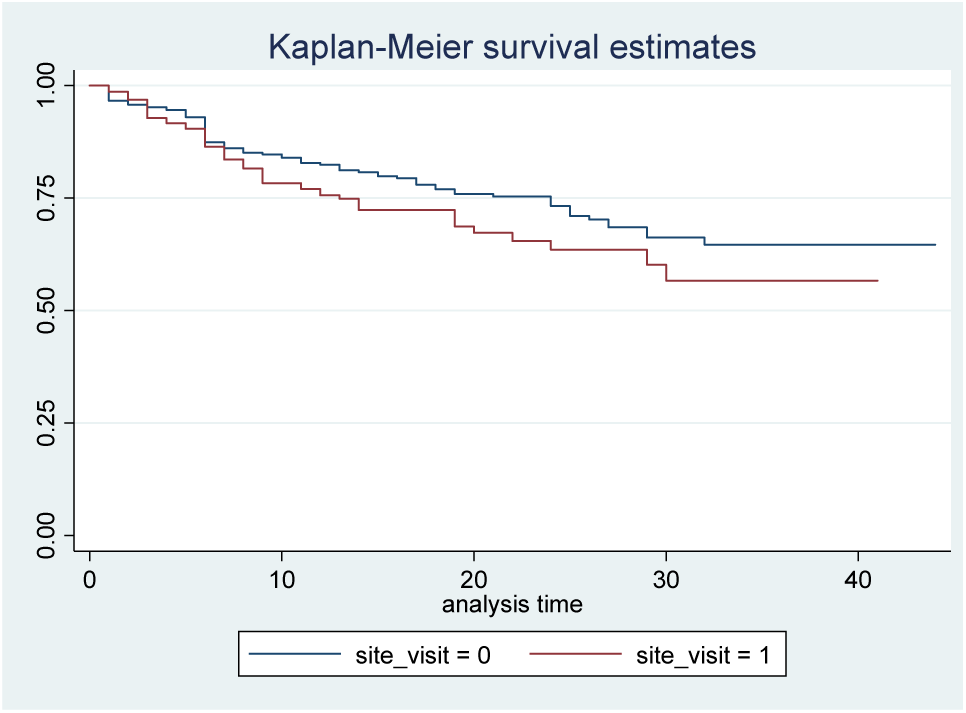
Continuation of postpartum modern reversible contraception by exposure to family planning information at service delivery points

The proportional hazard ratio is calculated using the Cox-proportional hazard model after adjusting for the relevant background variables. The results of the proportional hazard ratio analysis are presented in Table 6. Both the program variables have no or weak association with the continuation of postpartum contraception. For example, exposure to family planning (FP) information through health workers (Hazard ratio 0.882, p=0.476) is not significantly associated with the continuation of modern temporary methods. However, there is a weak association between exposure to FP information at service delivery points (Hazard ratio 1.336, p=0.100) and the continuation of modern temporary methods. This probably indicates that women who have contact with service delivery points have better access to contraceptive methods.

**Table 6.** Results of the proportional Hazard model

|  | Haz. Ratio | P>z |
| --- | --- | --- |
| Age in years | 0.982 | 0.509 |
| Children ever born (ref=1) |  |  |
| 2 | 0.775 | 0.199 |
| 3+ | 0.562 | 0.028 |
| Education in years | 0.960 | 0.045 |
| Muslim (Ref=Hindu) | 1.260 | 0.256 |
| Caste (Ref-SC/ST) |  |  |
| OBC | 0.832 | 0.345 |
| General | 0.964 | 0.900 |
| Engaged in paid work in last 12 months (ref=no) | 1.027 | 0.922 |
| Wealth quintiles (Ref = Quintile 1) |  |  |
| Quintile 2 | 0.945 | 0.807 |
| Quintile 3 | 0.895 | 0.664 |
| Quintile 4 | 1.237 | 0.461 |
| Quintile 5 | 1.130 | 0.718 |
| Inter-spousal communication regarding family planning (ref=no) <sup>1</sup> | 0.805 | 0.227 |
| Exposed to family planning messages in television (ref=no) | 0.984 | 0.925 |
| Index of contraceptive self-efficacy | 0.819 | 0.015 |
| Index of decision-making about the use of family planning | 0.905 | 0.209 |
| Exposure to family planning information through health workers | 0.882 | 0.476 |
| Exposure to family planning information at service delivery points | 1.336 | 0.100 |
| Note: ~ p<=0.10; *p<=0.05; **p<=0.01; ***p<=0.001 |  |  |
| <sup>1</sup> Women with missing information are not shown separately |  |  |

Similar to the findings from the adoption of postpartum reversible methods, the index of self-efficacy (Hazard ratio 0.817, p=0.015) continues to be associated with the continuation of postpartum modern reversible contraceptive methods. Specifically, a unit increase in the self-efficacy index reduces the risk of discontinuing modern reversible methods by 18%. The variables of parity and education significantly influenced the continuation of postpartum contraception. Women with a parity of three or higher (Hazard ratio 0.562, p=0.028) and education (Hazard ratio 0.960, p=0.045) were less likely to have discontinued postpartum contraception. Compared to women with one parity, women with parity three or higher are 44% less likely to have discontinued modern reversible methods. Regarding the education of women, which is the number of years of schooling completed, a year increase in the year of education reduces the risk of discontinuing postpartum contraception by four percent.

## Discussion

The study utilized data from the 2019 Output Track Survey, which surveyed 4,029 currently married women aged 15-49 residing in slums across 14 cities in three states of India. The analysis focused specifically on 1,503 women aged 15-34 who had given birth within the three years preceding the survey. The primary objective of the analysis was to examine the impact of program interventions on the adoption of postpartum contraception in these states.

In India, the increased institutional deliveries are an excellent opportunity for the health system to promote postpartum contraception. This opportunity continues over the extended postpartum period because the women would continue to be in touch with the health systems for postpartum care and child immunization (Moore et al. 2015; Cleland et al, 2015). Keeping this in view, the study finding that about half of the women were exposed to the program through health workers or visits to the health service delivery points is an encouraging step forward in reducing the unmet need for contraception. About 58 percent of women adopted postpartum contraception within six months, and it increased to 70 percent by 12 months of the postpartum period. The study also found that among the program exposure variables, exposure to family planning messages through health workers significantly increased the adoption of postpartum contraception in the study states. Previous studies also highlighted the importance of family planning discussions during the postpartum period in improving the adoption of contraceptive use after birth in resource-poor settings (Cleland et al, 2015; Mumah et al, 2015; Puri et al., 2021b; Puri et al., 2023; Asah-Opoku., 2023; Srivastava et al., 2022; Zavier and Santhya, 2013; Freeman-Spratt et al., 2023).

The study also highlighted the importance of women’s contraceptive self-efficacy and interspousal communication. The findings that women with contraceptive self-efficacy are more likely to adopt contraception and continue the use of contraception are also highlighted by other studies (Dhak et al., 2019; Ewerling et al., 2021; Singh et al., 2019). The absence of women’s agency acts as a deterrent for adolescents and youth in Bihar and Uttar Pradesh, hindering their adoption of contraception (Saggurti and Zavier, 2020a and 2020b).

The study also emphasized the significance of interspousal communication, as it positively influenced contraceptive usage. This underscores the importance of involving males in family planning efforts. The findings align with previous studies that have identified a lack of spousal communication as a barrier to accessing reproductive health services (Santhya and Jejeebhoy, 2015; Behera et al., 2016).

Findings also suggest that the one-year continuation rate of reversible modern contraceptive methods is approximately 75%. This rate of contraceptive continuation is similarly observed among postpartum IUD adopters in a study conducted in six states, where 68% continued for one year (Kumar et al., 2019), and a 78% twelve-month continuation rate was reported in Gujarat and Rajasthan (Singal et al., 2022). Evidence from the analysis of the continuation of modern reversible methods indicates that neither of the program variables was associated with adopting modern reversible methods. The study findings revealed that women’s education and high self-efficacy were significant factors contributing to the likelihood of continuing modern reversible methods. This suggests that a woman’s belief in her ability to manage and utilize contraceptives effectively can play a significant role in the continuation of these methods.

The study has some limitations. First, the limitations of the cross-section data analysis regarding causal association apply in the present analysis. Second, the study was designed to analyse overall contraceptive use. Therefore, the survey did not capture family planning-related interactions during the postpartum period. Finally, quality of care and postpartum visits, which are important in the adoption of postpartum contraception, were also not covered in the survey. Despite these limitations, the study findings are relevant in the context of the slum population.

The findings of the study have some programmatic implications. First, the health workers’ visit is essential in adopting postpartum contraception. At the same time, more studies are required to see why visits to the service delivery points did not show any association with the continuation of postpartum contraception. Secondly, once family planning is adopted, follow-up services are required so that the continuation of contraception will also increase. Third, the study also highlighted the importance of male involvement in family planning and life skills education programs for women. Finally, high-parity women must be motivated to adopt long-acting temporary or permanent methods.

## Data Availability

This study was quantitative in nature and we used a Stata data file for analysis. Authors will comply with journal's data policy and all data and related metadata underlying the findings reported in submitted manuscript will be deposited in an appropriate repository upon request from the journal. The datasets used for analysis are available from the first author/corresponding author.

## Acknowledgments

The authors acknowledge the TCI program team and the National Health Mission, Government of Uttar Pradesh, who supported the program’s design and implementation and gave permission for conducting data collection. We also thank the data collectors from Nielsen India Private Ltd and respondents of the output tracking surveys.

## Author contributions

ED, MKS, HS IS, KG, SB—TCI program design, questionnaire preparation, data collection; ED, AJFZ, MKS—conceptualized the paper, writing the first draft of the manuscript; ED, AJFZ, IS, NP—analysis, interpretation of results and manuscript preparation, finalized the manuscript. All authors reviewed and approved the final version of the manuscript.

